# Longitudinal dynamics of *Streptococcus pneumoniae* carriage and SARS-CoV-2 infection in households with children

**DOI:** 10.1101/2023.02.20.23286191

**Authors:** Willem R. Miellet, Rob Mariman, Dirk Eggink, Mioara A. Nicolaie, Janieke van Veldhuizen, Gerlinde Pluister, Lisa M. Kolodziej, Steven F.L. van Lelyveld, Sjoerd M. Euser, Elisabeth A.M. Sanders, Marianne A. van Houten, Krzysztof Trzciński

## Abstract

**Background:** To characterize interferences between *Streptococcus pneumoniae* and SARS-CoV-2 we investigated the longitudinal patterns of viral infection and pneumococcal carriage in households infected with SARS-CoV-2.

**Methods:** SARS-CoV-2 and pneumococcus were detected with quantitative molecular methods in saliva from members of eighty participating households. Samples were collected between October 2020 and January 2021 from n=197 adults and n=118 children of which n=176 adults and n=98 children had a complete set of ten samples collected within 42 days since enrolment. Time-dependent Cox models were used to evaluate the associations between SARS-CoV-2 and pneumococcal carriage.

**Results:** In the entire cohort, cumulative pneumococcal carriage and SARS-CoV-2 infection rates were 58% and 65%, respectively. Pneumococcal abundances were associated with an increased risk of SARS-CoV-2 infection (HR 1.14, 95% CI, 1.01 – 1.29, *P*=0.04) and delayed clearance of SARS-CoV-2 infection (HR 0.90, 95% CI, 0.82 – 0.99, *P*=0.03). Elevated viral loads were observed among pneumococcal carriers and individuals with high overall bacterial 16S abundances, however, there were no longitudinal differences in viral loads in linear mixed-effects models. Individuals with high 16S abundances displayed delayed viral clearance (HR 0.65, 95% CI 0.55 – 0.78, *P*<0.0001).

**Conclusions:** Although we found insufficient evidence for a strong impact of SARS-CoV-2 infection on pneumococcal carriage. Results from the current study suggest that pneumococcal carriers may have an increased risk of SARS-CoV-2 infection and high pneumococcal abundances and 16S abundances may be associated with elevated viral loads and delayed clearance of SARS-CoV-2 infection.

## INTRODUCTION

Epidemiological studies and animal co-infection models suggest strong synergistic interactions between certain respiratory viruses and *Streptococcus pneumoniae* [1]. For one, influenza virus, respiratory syncytial virus, and rhinovirus provoke host immune responses that can disturb colonizing resistance induced by the microbiota of the upper respiratory tract (URT), mute innate immune recognition of pneumococcus and increase the risk of secondary pneumococcal pneumonia [2-8]. Concomitantly, inflammation associated with pneumococcal colonization may facilitate viral respiratory infections [9-12].

Relatively little is known about interferences between pneumococcus and SARS-CoV-2. However, between 2020 and 2021 multiple countries reported declines in incidence of invasive pneumococcal disease (IPD) [13-15] and co-infections with pneumococcus were rare in COVID-19 patients [14, 16]. Although high rates of empiric antibiotic therapy in severe COVID-19 cases could have diminished pneumococcal disease incidence and contribute to low co-infection rates [17-19], the decline in IPD was primarily attributed to non-pharmaceutical interventions (NPIs) [13, 14]. It has been assumed that NPIs that included social distancing, wearing facemasks, closure of schools, restaurants, shops and cultural facilities and the advice to work from home limited pneumococcal transmission, thereby reducing the prevalence of pneumococcal carriage and consequently the incidence of IPD during pandemic [13]. However, several studies have reported that pneumococcal carriage rates in children were relatively unaffected during the pandemic when compared with the pre-COVID-19 period [15, 20]. Danino *et al*. concluded that declines in pneumococcal disease were likely driven by declines in seasonal respiratory virus circulation and not by reduced pneumococcal transmission [15].

URT inflammation associated with pneumococcal colonization may facilitate propagation of viral infection in the airways of exposed individuals [9-11]. While few clinical studies have investigated co-infections between SARS-CoV-2 and pneumococcus, none of the published papers have reported on possible interference between the two pathogens within households and studied longitudinal effects. To investigate the effect of pneumococcal colonization on SARS-CoV-2 infection and *vice versa*, we have conducted a longitudinal household study using molecular diagnostic methods to detect SARS-CoV-2 and pneumococcus in self-collected saliva samples [6, 21]. In our study, we found insufficient evidence for a strong impact of SARS-CoV-2 infection on pneumococcal carriage. However, pneumococcal carriers, children in particular, exhibited an increased risk of SARS-CoV-2 infection. High pneumococcal abundances and 16S abundances were associated with elevated viral loads and delayed clearance of SARS-CoV-2 infection.

## MATERIALS and METHODS

### Study Design and Ethics statement

We investigated pneumococcal carriage in a prospective observational cohort study [21] conducted between October 2020 and January 2021. Households were included in the study when a household member below 65 years of age tested positive for SARS-CoV-2 using qPCR within 72 hours prior to inclusion and at least 2 other household members consented to participate in the study. Enrolled study participants were asked to self-collect saliva samples over a period of six weeks. Written informed consent was obtained from all study participants or from their legal guardians. The study was reviewed and approved by the Medical Ethical Committee of the Vrije Universiteit university Medical Centre (Vumc), The Netherlands (reference nr. 020.436).

### Sample Collection

As previously described by Kolodziej *et al*., saliva samples were collected on day 1, 3, 5, 7, 10, 14, 21, 28, 35, and 42 since inclusion [21]. In short, individuals aged ≥5 years were asked to collect saliva, by spitting approximately 2 ml into a Genefix Saliva Collection device without buffer (Isohelix). From individuals aged <5 years saliva was collected with two Oracol (S10) sponges (Malvern Medical Developments). Samples were stored by participants in their home freezer and within three weeks transported on dry ice to the diagnostic laboratory for storage at -80°C.

### SARS-CoV-2 and *S. pneumoniae* detection with quantitative molecular methods

Nucleic acids were extracted from 200 µl of saliva using a MagNApure 96 system (Roche) with Equine arteritis virus and yeast tRNA added as internal control and stabilizer, respectively. Nucleic acids were eluted into 50 µl Tris EDTA buffer. To detect SARS-CoV-2, 5 *μ*l of the eluate was tested in reverse-transcriptase qPCR with primers and probe targeting the SARS-like beta coronavirus-specific *Egene* [22]. Pneumococcal DNA was detected by testing 5.5 *μ*l of a sample in single-plex qPCRs with primers and probes targeting DNA sequences within genes encoding for pneumococcal iron uptake ABC transporter lipoprotein (PiaB) [23], and major pneumococcal autolysin (LytA) [24] as previously described [25]. We quantified 16S with qPCR as a marker of overall bacterial abundance [6]. Molecular detection of pneumococcal serotypes is described in the **Supplement**.

### Statistical Analysis

Data analysis was performed in R version 4.2.2. Samples were considered positive by qPCR for *piaB, lytA* and *Egene* when C_q_s were below 40. For qPCR-based serotyping results concordance with *piaB/lytA* was required [26]. Intraclass correlations (ICCs (3,1)) were calculated in a two-way mixed-effects model with consistency relationship (*piaB* C_q_ = *lytA* C_q_ + systematic error) and of single measurement type [27], and Bland-Altman plots [28] were used to assess agreement between qPCR measurements using the “ïrr” and “blandr” R packages, respectively. Unpaired and paired carriage rates were compared with Fisher’s exact test and McNemar’s test, respectively. Relative pneumococcal abundances were calculated by dividing pneumococcal abundances by overall bacterial abundances (16S). Bacterial and pneumococcal abundances were compared by means of a permutation test equivalent of Mann-Whitney U test with blocking by age group [29]. Linear mixed-effects modeling was conducted to assess the association between SARS-CoV-2 infection and pneumococcal carriage status, and between SARS-CoV-2 infection and 16S abundances, where individuals at time 1 were classified as having low (< median at time 1) or high (≥ median at time 1) 16S abundance. A random intercept was used in linear-mixed effects model to account for longitudinal intra-individual variance, and to address inter-individual variance. The time to SARS-CoV-2 infection and time to SARS-CoV-2 infection clearance in relationship to pneumococcal carriage, pneumococcal abundance and log-transformed overall bacterial (16S) abundance were assessed by means of stratified time-dependent Cox proportional hazards models, where strata were defined by age groups. In order to estimate age-specific hazard rates (HR), two age groups were considered, children (<18 years) and adults (≥ 18 years). The proportionality assumption was tested using the Schoenfeld residuals test. Model selection was performed by means of the Akaike information criterion and model checking was done by comparing the model-based survival curves with the Kaplan-Meier estimator. A *p* value of <0.05 was considered significant.

## RESULTS

Of 337 individuals from whom saliva was collected, 11 had incomplete questionnaire data and 52 fewer than ten samples collected. As such, analysis was limited to 274 (81.3.%) individuals of 80 households (**Table 1)**. Sixty-five percent of individuals (177/274) and 95% (76/80) of households tested positive for SARS-CoV-2 infection in saliva during the six weeks of screening [21]. Per study design, almost all SARS-CoV-2 infections occurred within the first 10 days since enrolment and the number of SARS-CoV-2 infected individuals began to decline after the first 14 days of the study period [21].

**Table 1:** Study participant characteristics.

| Group | Overall<br>(n=274) | SARS-CoV-2<br>infected<br>(n=177) | SARS-CoV-2<br>uninfected<br>(n=97) | *S. pneumoniae*<br>carriers<br>(n=160) | *S. pneumoniae*<br>noncarriers<br>(n=114) | Co-infected<br>(n=106) |
| --- | --- | --- | --- | --- | --- | --- |
| median age, years (range) | 32 (0-64) | 32 (0-63) | 30 (1-64) | 17 (1-63) | 42 (0-64) | 22 (1-63) |
| age group, n (%) |  |  |  |  |  |  |
| < 12 | 51 (18.6) | 29 (16.4) | 22 (22.7) | 45 (28.4) | 6 (5.3) | 25 (23.6) |
| 12-17 | 47 (17.2) | 31 (17.5) | 16 (16.5) | 36 (22.5) | 11 (9.6) | 25 (23.6) |
| 18-39 | 65 (23.7) | 49 (27.7) | 16 (16.5) | 31 (19.4) | 34 (29.8) | 25 (23.6) |
| 40-49 | 74 (27.0) | 44 (24.9) | 30 (30.9) | 35 (21.9) | 39 (34.2) | 21 (19.8) |
| 50-65 | 37 (13.5) | 24 (13.6) | 13 (13.4) | 13 (8.1) | 24 (21.1) | 10 (9.4) |
| sex, n (%) |  |  |  |  |  |  |
| male | 135 (49.3) | 87 (49.2) | 48 (49.5) | 85 (53.1) | 50 (43.9) | 56 (52.8) |
| female | 139 (50.7) | 90 (50.8) | 49 (50.5) | 75 (46.9) | 64 (56.1) | 50 (47.2) |
| comorbidities, n (%) | 30 (10.9) | 14 (7.9) | 16 (16.5) | 14 (8.8) | 16 (14.0) | 7 (6.6) |
| symptomatic, n (%) | 167 (60.9) | 139 (78.5) | 28 (28.9) | 96 (60.0) | 71 (62.3) | 78 (73.6) |
| median household size, n (range) | 4 (3-6) | 4 (3-6) | 4 (3-6) | 4 (3-6) | 4 (3-6) | 4 (3-6) |
| presence of a child aged <5 years<br>in a household, n (%) | 54 (19.7) | 37 (20.9) | 17 (17.5) | 48 (30.0) | 6 (5.3) | 31 (29.2) |

### Pneumococcal Carriage Prevalence Rates in Households with Individuals Infected with SARS-CoV-2

Altogether, 160 individuals (58% of 274) tested positive for pneumococcus by qPCR. This corresponded to 40% (1083/2740) of samples positive for pneumococcus using a dual-target approach with *piaB* and *lytA* genes, with significant correlations between *piaB* and *lytA* C_q_s (**Fig. 1**) and ICCs of 0.89 (95% CI 0.87 – 0.90) indicative of good reliability. Similar ICCs were observed at each of the ten sampling events separately (**Fig. S1**). During a follow-up of six weeks cumulative pneumococcal carriage and SARS-CoV-2 infection rates were 58.4% (160 of 274) and 64.6% (177 of 274), respectively. We observed a mean pneumococcal carriage rate of 35.8% (98/274, 95% CI 30-42%) (**Table S1**). The proportion of pneumococcal carriers with SARS-CoV-2 infection was high during the first two weeks of the study with on average 54.1% of pneumococcal carriers concurrently positive for SARS-CoV-2 and a cumulative co-infection rate of 33.9% (93 of 274). Despite high co-infection rates, pneumococcal carriage rates remained relatively constant throughout the study period (**Fig. 2**), suggesting little impact of SARS-CoV-2 infection on pneumococcal carriage rates. Rates of pneumococcal carriage were highest among children <12 years of age (88.2%, **Table 1**) and households with young children (<5 years; n=15) displayed significantly increased rates of pneumococcal carriage compared with other households (Fisher’s exact test, *P*<0.0001, OR 7.7 [95% CI 3.1-22.8]).

**Figure 1:**
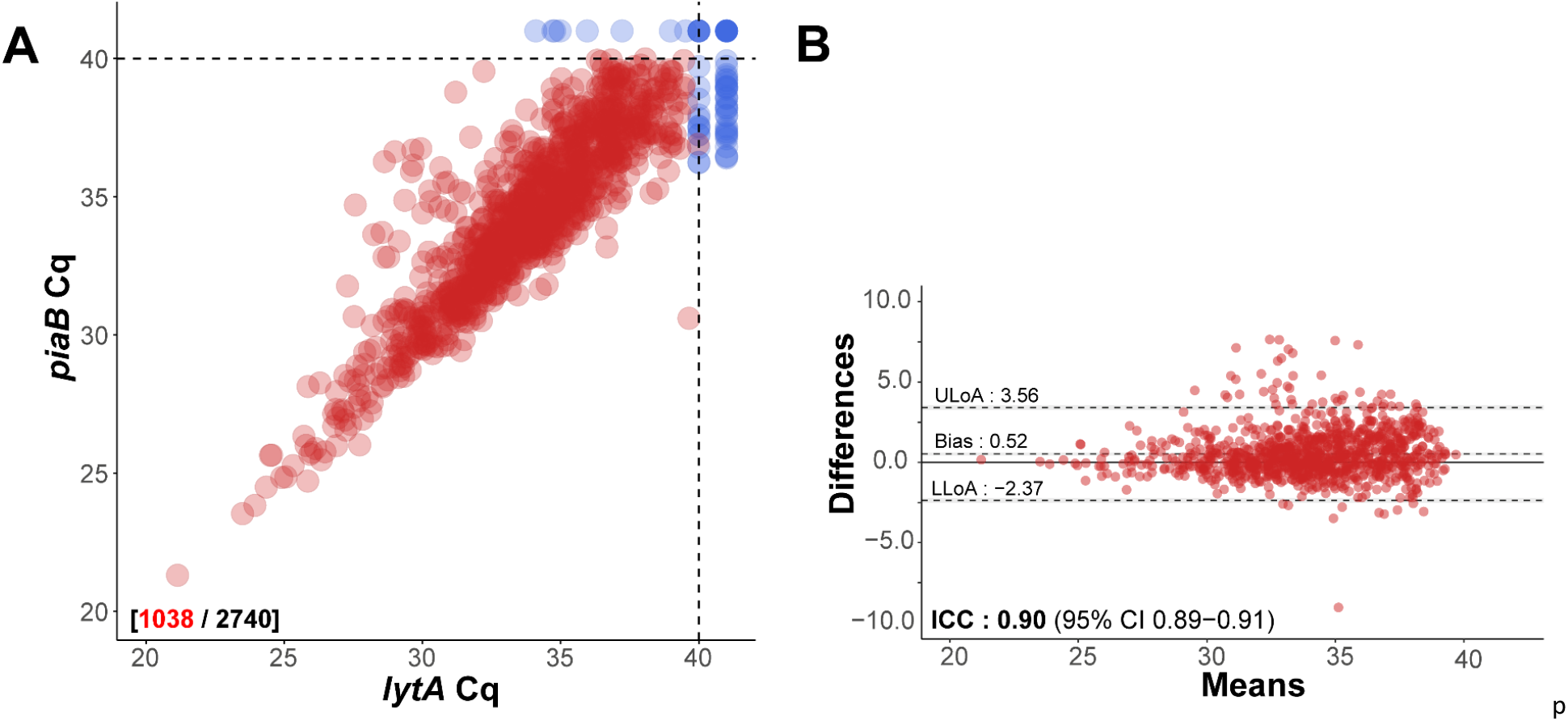
(A) Scatter plot displaying C_q_ piaB and lytA *quantification* for all individuals (n=274) and all samples (n=2740). (B) Bland-Altman plot displaying the extent of agreement in quantitative C_q_ measur*e*ments for *piaB* and *lytA* among positive samples. Samples were classified as positive when measurements for *piaB* and lytA were both <40 C_q_ (indicated by dashed lines). Red and blue dots represent samples positive and negative for *S. pneumoniae*, respectively. Numbers coloured red indicate the number of samples classified as positive for pneumococcus and black. ICC: intraclass correlation coefficient - a single score intraclass correlation coefficient with two-way model (consistency) was used for reliability analysis. The Bland-Altman plot illustrates the degree of agreement between *piaB* and *lytA* C_q_ measurements among positive samples. The mean difference in measurements is indicated by a dashed grey line and the standard deviations of the mean, upper limit of agreement (ULoA), and lower limit of agreement (LLoA) are also shown. In this Bland-Altman plot differences above the line of equality, indicated by a solid line, are for samples of which *lytA* C_q_s where lower than *piaB* C_q_s. ICC values <0.50, 0.50-0.75, 0.75-0.90 and >0.90 are indicative of poor, moderate, good, and excellent reliability, respectively.

**Figure 2:**
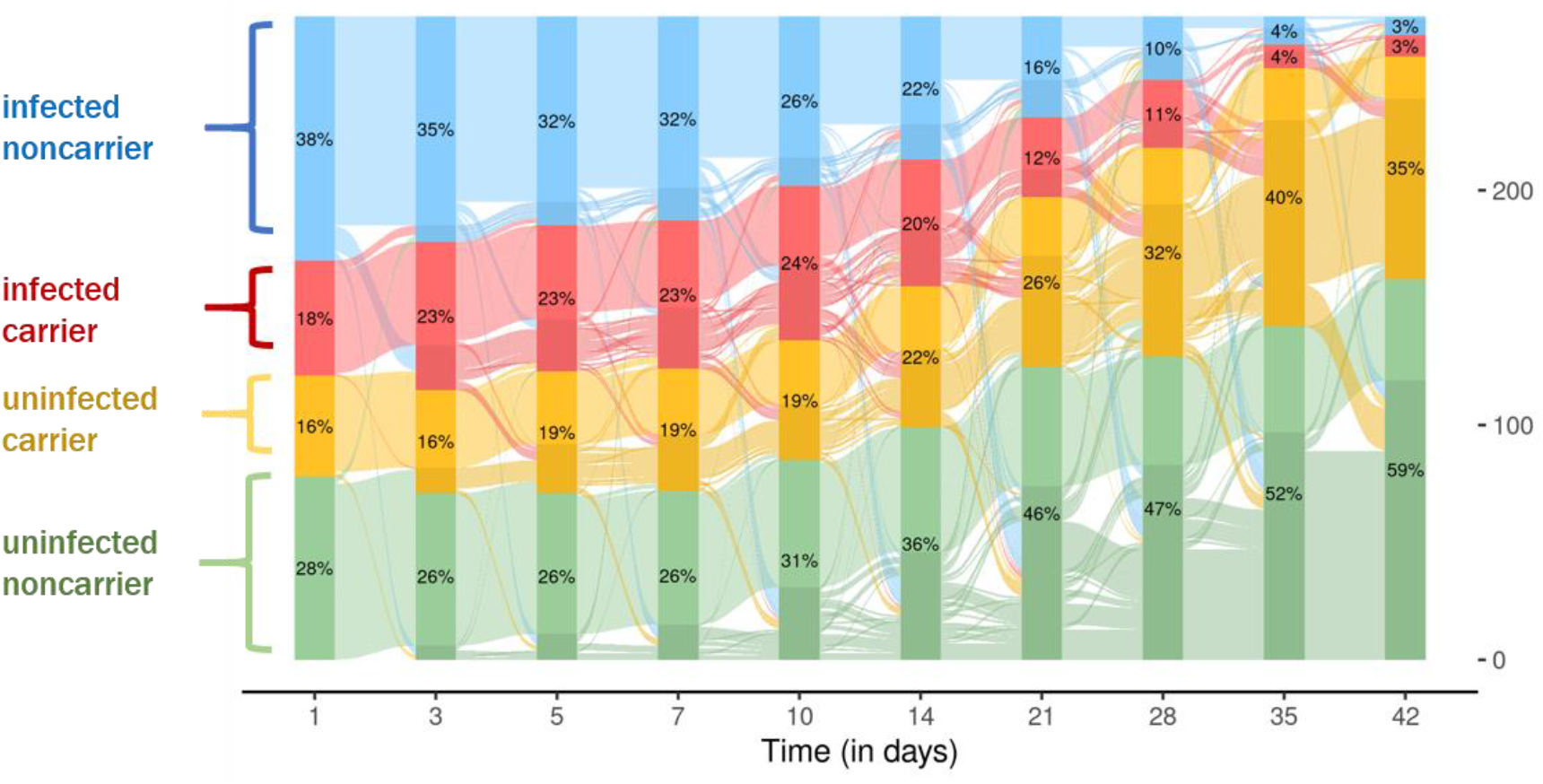
Alluvial diagram displaying longitudinal changes across the study period in participant status for SARS-CoV-2 infection and pneumococcal carriage among n=274 participants. Infected noncarriers, infected carriers, uninfected carriers, uninfected noncarriers are colored in blue, red, yellow and green, respectively. Percentages of groups are shown in bars. The right y-axis indicates the number of individuals.

### Hazards Model Analysis

We sought to evaluate the relationship between pneumococcal carriage and SARS-CoV-2 infection. To this goal, only samples from individuals who tested negative at enrolment were considered (n=121 individuals experiencing n=23 virus acquisition events). We fit a Cox proportional hazards model to the time to SARS-CoV-2 infection. In a multivariate model, pneumococcal abundances and 16S abundances were used as covariates. To account for events within the same family, we calculated robust standard errors for effects sizes. Next, we performed model selection (**Table S2, Fig. S2**). Using the multivariable model, pneumococcal abundances were associated with an increased risk for SARS-CoV-2 infection (HR 1.14, 95% CI, 1.01 – 1.29), whereas we did not find sufficient evidence for an association between 16S abundances and SARS-CoV-2 infection (HR 0.86, 95% CI, 0.58 – 1.27). The cumulative hazard rates across the study period were higher for children (HR 2.14) than for adults (HR 0.83), suggesting that the relationship between pneumococcal abundances and SARS-CoV-2 infection was primarily associated with children.

Next, we assessed the association between pneumococcal carriage and SARS-CoV-2 infection clearance. We therefore limited the analysis to samples from individuals who tested positive for SARS-CoV-2 at enrolment (n=153 individuals experiencing n=134 events) and we used stratified Cox regression analysis to model the time to SARS-CoV-2 infection clearance. Multivariate models that included covariates for pneumococcal abundance and 16S abundance, were considered for model selection (**Table S3, Fig. S3**). In the multivariable model both pneumococcal abundances (HR 0.90 95% CI, 0.82 – 0.99) and 16S abundances (HR 0.68 95% CI, 0.55 – 0.78) before day 21 were significantly associated with a decreased risk for SARS-CoV-2 infection clearance (**Table S3**).

### Pneumococcal abundances among SARS-CoV-2 infected individuals

We then sought to determine whether SARS-CoV-2 infection impacted pneumococcal abundances and bacterial presence in the upper respiratory airways. SARS-CoV-2 infected individuals exhibited significantly elevated pneumococcal abundances at day 10, 21 and 28 when compared with uninfected individuals (**Fig. S4A**). Likewise, significantly elevated 16S abundances among SARS-CoV-2 infected individuals were observed at day 10, 14, 21 and 28 (**Fig. S4B**). However, no significant differences between SARS-CoV-2 infected and uninfected individuals were observed for pneumococcal relative abundances (**Fig. S4C**). In line with this, we did not observe considerable shifts in serotype carriage composition across the study period (**Fig. S5**).

### Increased viral loads in individuals carrying pneumococcus

Thereafter, we quantified *Egene* as a marker of SARS-CoV-2 viral loads from individuals who tested positive for SARS-CoV-2 at enrolment and compared viral loads among pneumococcal carriers and noncarriers, and individuals with high or low 16S abundances. Significantly elevated viral loads were observed at multiple sampling time points for pneumococcal carriers when compared with noncarriers (**Fig. 3A**). Individuals with high 16S abundances also displayed significantly increased viral loads at multiple sampling events when compared with individuals with low 16S abundances (**Fig. 3B**). We subsequently modelled longitudinal trajectories of log-transformed viral loads using linear mixed-effects modelling. Trajectories were estimated from positive samples by censoring measurements with *Egene* C_q_s 40 or higher. We did not observe significant differences in longitudinal viral load trajectories between pneumococcal carriers versus noncarriers (**Fig. S6A-B**), and between individuals with high versus low 16S abundances (**Fig. S6C-D**).

**Figure 3:**
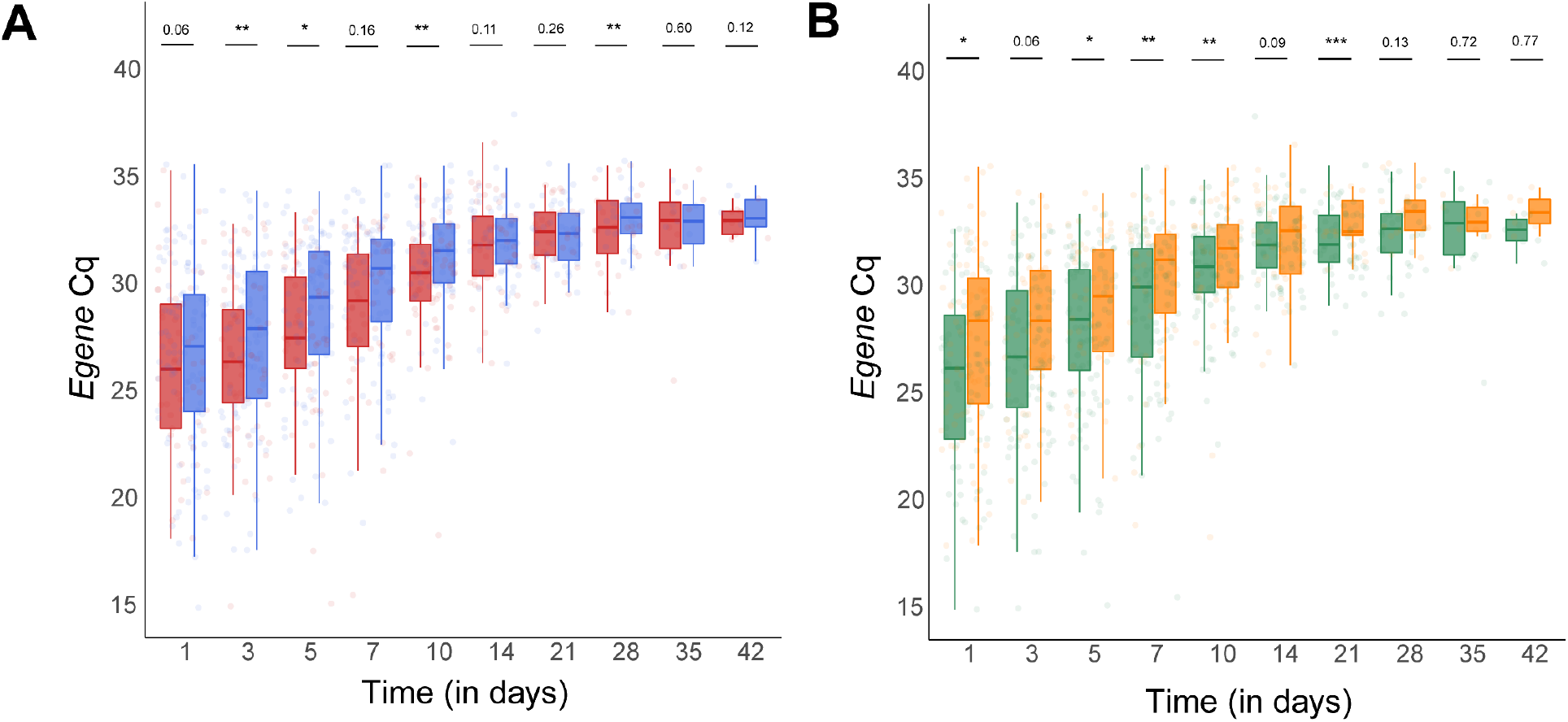
Boxplots of SARS-CoV-2 viral loads from individuals positive for SARS-CoV-2 since study onset (t=1) and stratified by pneumococcal carriage status (A) and overall bacterial abundance (B). Viral loads were compared blocked by age group (< 18 years old or ≥ 18 years old)with a permutation test equivalent of Mann-Whitney U test. Pneumococcal carriers display significantly increased viral loads at day 3, 5, 10 and 28 of study period when compared with noncarriers. Individuals with increased bacterial abundance display significantly increased viral loads at day 1, 5, 7, 10 and 21 of study period when compared with individuals with reduced bacterial abundance. Medians are indicated by a solid line in a boxplot.

## DISCUSSION

Interplay between respiratory viruses and pneumococcus are often synergistic drivers of respiratory infections [8, 30]. In the current study, we investigated whether associations exist between SARS-CoV-2 infection and pneumococcal carriage among children and non-elderly adults.

We observed high rates of pneumococcal carriage within households during nationwide implementation of NPIs in the Netherlands in 2020/2021 [31]. Despite high coinfection rates we have found insufficient evidence for a strong impact of SARS-CoV-2 infection on pneumococcal carriage rates and pneumococcal abundances. However, pneumococcal carriers exhibited an increased risk of SARS-CoV-2 infection and delayed viral clearance. Delayed viral clearance was also observed among individuals with high 16S abundances yet we observed no association between 16S abundances and SARS-CoV-2 infection rates. Both pneumococcal carriers and individuals with high 16S abundances exhibited increased SARS-CoV-2 viral loads at multiple sampling events. However, differences in longitudinal viral load trajectories among pneumococcal carriers or individuals with high 16S abundances were not significant.

Despite the imposition of NPIs and a decline in seasonal respiratory virus circulation during the study period [32] there was a substantial presence of pneumococcal carriage. Our observations are in line with reports by others [15, 20], suggesting that pneumococcal carriage prevalence rates did not decline during the COVID-19 pandemic and during NPIs. In contrast, the incidence of IPD dropped sharply during the 2020/2021 influenza season [13-15]. Danino *et al*. have attributed this decline in IPD to a drop in seasonal respiratory virus circulation following the imposition of NPIs and travel restrictions across countries [15].

Pneumococcal carriage rates were relatively constant throughout the study period. SARS-CoV-2 infected individuals exhibited elevated pneumococcal abundances and 16S abundances at several time points. However, no significant differences between SARS-CoV-2 infected and uninfected individuals were observed for relative pneumococcal abundances. Moreover, we have observed no substantial shift in sample serotype composition during the study period. Collectively, these observations suggest no marked impact of SARS-CoV-2 infection unique to pneumococcal colonization. Our findings are in line with studies describing that co-infections of SARS-CoV-2 and pneumococcus are uncommon [17, 33].

We performed Cox regression analysis to study possible interactions between SARS-CoV-2 and pneumococcus. As such, we modelled the impact of pneumococcal abundances independently from 16S abundances on SARS-CoV-2 infection rates and on SARS-CoV-2 infection clearance rates. Pneumococcal abundances were associated with an increased risk for SARS-CoV-2 infection, while there was insufficient evidence for an altered risk among individuals with high 16S abundances. High cumulative hazard rates among children but not among adults indicated that the relationship between pneumococcal abundances and SARS-CoV-2 infection was primarily associated with children.

In an observational study among older adults vaccination with PCV13 was reported to the reduce the risk of COVID-19 diagnosis, COVID-19 hospitalization and fatal COVID-19 outcome [17], suggesting that PCV13 vaccine-type pneumococcal carriage could predispose older adults to adverse outcomes associated with COVID-19. Another study among adults reported an increased odds of SARS-CoV-2 infection among pneumococcus-positive adults attending a COVID-19 testing clinic, yet in the same study pneumococcus-positive adults attending an outreach facility did not exhibit an increased odds of SARS-CoV-2 infection [34], suggesting that the association between pneumococcal carriage and SARS-CoV-2 may be non-causal. While previous studies focused on older individuals [17, 34], we investigated possible viral-bacterial interactions in children and non-elderly adults. Although we find that pneumococcal abundances were associated with increased SARS-CoV-2 infection rates, due to the limit number of events we cannot exclude that the reported association may have been attributable to confounding factors.

Findings from a study in the United Kingdom have suggested that IgA and CD4^+^ T-cell responses to SARS-CoV-2 may be impaired among pneumococcal carriers [35]. Of note, CD4^+^ T-cell mediated responses are also critical for clearance of pneumococcal colonization. Impairment of immune responses to SARS-Cov-2 also prolongs viral infection [36]. Consequently, one would expect delayed clearance of SARS-CoV-2 infection among pneumococcal carriers. Cox regression analysis of SARS-CoV-2 infection clearance rates indicated that high pneumococcal abundances were indeed associated with delayed clearance of SARS-CoV-2 infection. However, since high 16S abundances were also associated with delayed viral clearance, we do not find an association unique to pneumococcus.

High 16S abundances of the URT have previously been linked to dysbiosis of the URT microbiota in adults with community-acquired pneumonia [37]. The observed association between increased 16S abundances and delayed SARS-CoV-2 infection clearance in this study may therefore result from COVID-19 induced URT microbiome dysbiosis. Our observation is also in line with a study describing distinct URT microbiome composition profiles among hospitalized COVID-19 patients when compared with healthy individuals, which were linked to URT microbiota dysbiosis and were shown to correlate with COVID-19 severity [38].

Among study participants infected with SARS-CoV-2 at enrolment, pneumococcal carriers and individuals with high 16S abundances exhibited elevated SARS-CoV-2 viral loads at multiple sampling events, suggesting that presence of pneumococcal carriage could have impact SARS-CoV-2 infection progression. Modelling of longitudinal viral load patterns indicated that pneumococcal carriage was not associated with altered viral load dynamics. Since elevated SARS-CoV-2 viral loads were observed among pneumococcal carriers and individuals with high 16S abundances, it appears likely that these findings are also related to COVID-19 induced URT microbiota dysbiosis rather than exacerbation of SARS-CoV-2 infection due to pneumococcal colonization. Of note, since this study was conducted at a time prior to the emergence of SARS-CoV-2 variants of concern (VOCs) in the Netherlands, interferences we observed could be affected by higher transmissibility of VOCs [31].

As this study was primarily designed to evaluate the use of saliva in monitoring SARS-CoV-2 and to study SARS-CoV-2 household transmission dynamics [21], it has number of limitations for analysis of pneumococcal carriage and interactions between pneumococcus and SARS-CoV-2. The age group at greatest risk for both pneumococcal disease and COVID-19 was not included in this study, as such interactions between pneumococcus and SARS-CoV-2 specific for older age have been missed. The number of secondary cases of SARS-CoV-2 infections after the first sampling event were low, limiting the statistical power available for Cox regression analysis of SARS-CoV-2 infection. Moreover, the overall study size limited our ability to adjust for potential confounding factors in Cox regression analysis. Pneumococcal carriage episodes may have been missed due to the omission of a culture-enrichment procedure for pneumococcal detection [39].

In conclusion, in a cohort of children and non-elderly adults we observed high rates of pneumococcal carriage and coinfections with SARS-CoV-2. While we find insufficient evidence for a strong impact of SARS-CoV-2 infection on pneumococcal carriage rates and pneumococcal abundances, we do observe an increased risk of SARS-CoV-2 infection among pneumococcal carriers, children in particular. Moreover, individuals with high pneumococcal abundances or 16S abundances exhibited delayed SARS-CoV-2 infection clearance and elevated viral loads, suggesting dysbiosis of the URT microbiome following SARS-CoV-2 infection.

## Data Availability

All data produced in the present study are available upon reasonable request to the authors.

## AUTHOR CONTRIBUTIONS

W.R.M., E.A.M.S., M.A.v.H. and K. T. contributed to the conception and design of the study. D.E., J.v.V., G.P., L.M.K., S.v.L., S.M.E. and R.M. participated in acquisition of data. R.M., D.E., S.v.L. and S.M.E. coordinated the laboratory analyses. W.R.M., R.M., M.A.N., J.v.V., L.M.K. and K.T. were responsible for data analyses and interpretation. W.R.M., M.A.N., L.M.K. and J.v.V. verified the underlying data W.R.M., R.M. and K.T. wrote the manuscript. All authors reviewed and approved the final version of the manuscript.

## ACKNOWLEDGEMENTS

We thank all the participants for their contribution to the study. We thank the team of the Public Health Services Kennemerland, for providing information to the (potential) participants; the Regional Public Health Laboratory (Streeklab) Kennemerland for laboratory analyses; and the research team of the Spaarne Gasthuis Academy, particularly Greetje van Asselt, Jacqueline Zonneveld, Sandra Kaamer van Hoegee, and Mara van Roermund for their hard work and Coen Lap for his efforts concerning the continuation of the SARSLIVA study. We thank the laboratory team from the virology department at National Institute for Public Health and the Environment (RIVM). We thank Tessa Nieuwenhuijsen for her assistance in the laboratory.

---

**Table S1:** Rates across study observation period.

| Group | T=1 | T=3 | T=5 | T=7 | T=10 | T=14 | T=21 | T=28 | T=35 | T=42 |
| --- | --- | --- | --- | --- | --- | --- | --- | --- | --- | --- |
| SARS-CoV-2 infection, n (%) | 153 | 159 | 151 | 150 | 138 | 115 | 77 | 56 | 22 | 17 |
|  | (55.8) | (58.0) | (55.1) | (54.7) | (50.4) | (42.0) | (28.1) | (20.4) | (8.0) | (6.2) |
| pneumococcal carriage, n (%) | 92 | 107 | 114 | 115 | 117 | 114 | 106 | 118 | 120 | 104 |
|  | (33.6) | (39.1) | (41.6) | (42.0) | (42.7) | (41.6) | (38.7) | (43.1) | (43.8) | (38.0) |

Observed SARS-CoV-2 infection and *S. pneumoniae* carriage rates across the six week study period among 274 individuals, including 98 children and 176 non-elderly adults.

**Table S2:** SARS-CoV-2 infection acquisition model selection.

| Model | Coefficient (95% CI) | P-value | AIC |
| --- | --- | --- | --- |
| Model I: |  |  | 188.13 |
| log-transformed *piaB* abundance | 1.14 (0.99 – 1.31) | 0.065 |  |
| Model II: |  |  | 191.10 |
| log-transformed 16S abundance | 0.87 (0.59 – 1.29) | 0.487 |  |
| Model III: |  |  | 189.55 |
| log-transformed *piaB* abundance | 1.14 (0.99 – 1.31) | 0.096 |  |
| log-transformed 16S abundance | 0.86 (0.57 – 1.27) | 0.686 |  |
| Model IV: |  |  | 189.55 |
| log-transformed *piaB* abundance | 1.14 (1.01 – 1.29) | 0.038 |  |
| log-transformed 16S abundance | 0.86 (0.58 – 1.27) | 0.673 |  |

AIC: Akaike information criterion, Number of included events were n=24.

**Table S3:**
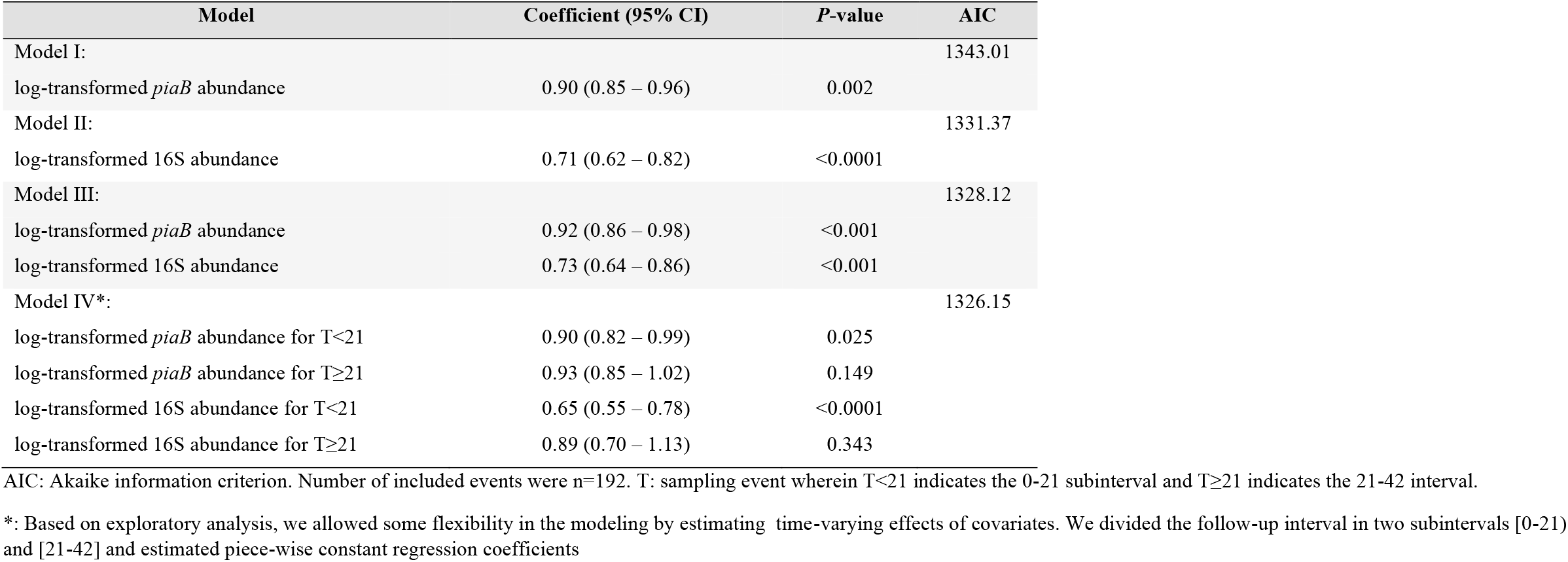
SARS-CoV-2 infection clearance model selection.

| Model | Coefficient (95% CI) | P-value | AIC |
| --- | --- | --- | --- |
| Model I: |  |  | 1343.01 |
| log-transformed *piaB* abundance | 0.90 (0.85 – 0.96) | 0.002 |  |
| Model II: |  |  | 1331.37 |
| log-transformed 16S abundance | 0.71 (0.62 – 0.82) | <0.0001 |  |
| Model III: |  |  | 1328.12 |
| log-transformed *piaB* abundance | 0.92 (0.86 – 0.98) | <0.001 |  |
| log-transformed 16S abundance | 0.73 (0.64 – 0.86) | <0.001 |  |
| Model IV*: |  |  | 1326.15 |
| log-transformed *piaB* abundance for T<21 | 0.90 (0.82 – 0.99) | 0.025 |  |
| log-transformed *piaB* abundance for T≥21 | 0.93 (0.85 – 1.02) | 0.149 |  |
| log-transformed 16S abundance for T<21 | 0.65 (0.55 – 0.78) | <0.0001 |  |
| log-transformed 16S abundance for T≥21 | 0.89 (0.70 – 1.13) | 0.343 |  |

AIC: Akaike information criterion. Number of included events were n=192. T: sampling event wherein T<21 indicates the 0-21 subinterval and T≥21 indicates the 21-42 interval.
\*: Based on exploratory analysis, we allowed some flexibility in the modeling by estimating time-varying effects of covariates. We divided the follow-up interval in two subintervals [0-21) and [21-42] and estimated piece-wise constant regression coefficients

**Figure S1:**
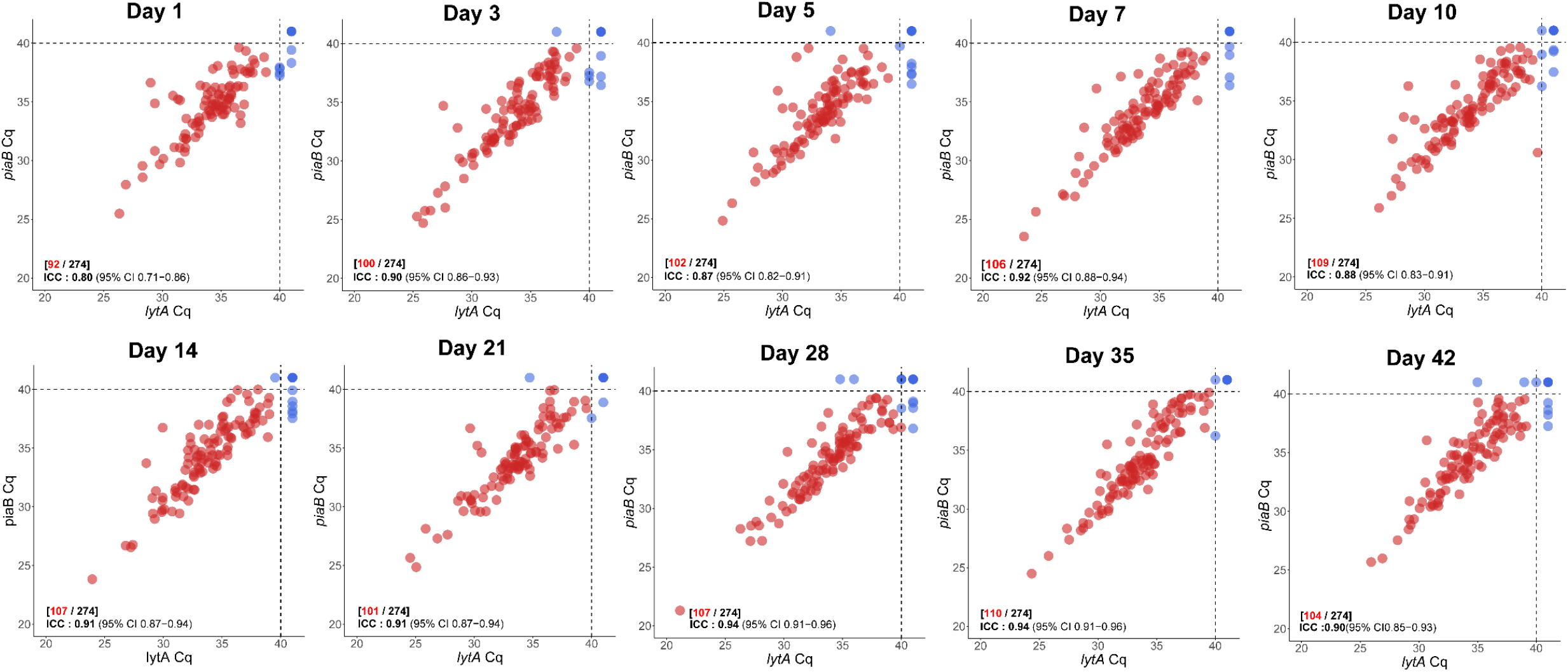
Scatter plot displaying C_q_ measurements for *piaB* and *lytA* targets for all individuals at individual sampling events. Red and blue dots represent samples positive and negative for *S. pneumoniae*, respectively. Samples were considered to be positive for *S. pneumoniae* when Cq for both *piaB* and *lytA* were below 40 C_q_, the C_q_ threshold is indicated by dashed lines. Numbers coloured red indicate the number of samples classified as positive for pneumococcus and black. ICC: intraclass correlation coefficient - a single score intraclass correlation coefficient with two-way model (consistency) was used for reliability analysis. ICC values <0.50, 0.50-0.75, 0.75-0.90 and >0.90 are indicative of poor, moderate, good, and excellent reliability, respectively.

**Figure S2:**
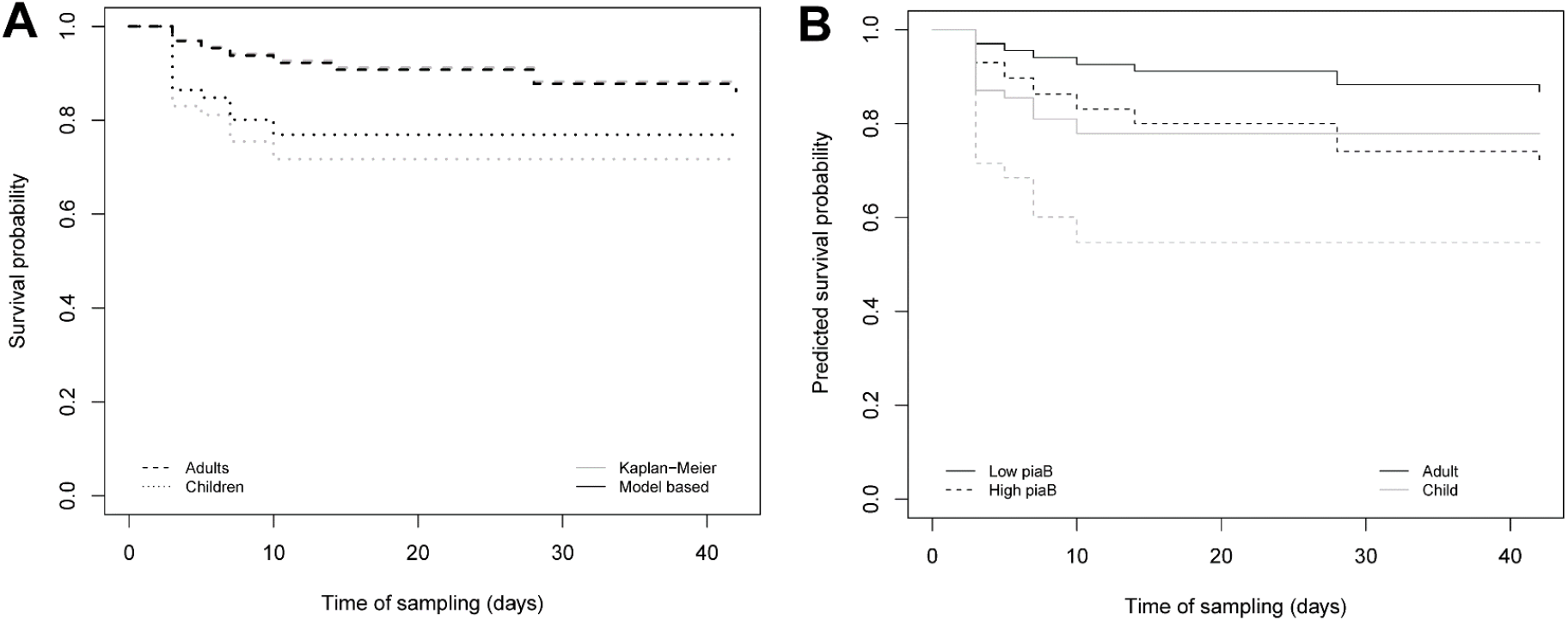
**(A)** The survival probability of time to infection acquisition estimated by the non-parametric Kaplan-Meier estimator (in grey) and model-based (in black) and the respective 95% confidence intervals. While the two survival curves for children are virtually the same up to day 5, after day 5 the model-based curve slightly overestimates the non-parametric estimator. This difference is most likely due to the uncertainty of estimation caused by the reduced number of events after day 5 in our sample. **(B)** Predicted survival probabilities based on the Cox proportional hazards model (model IV of Table S2), stratified by age group, were compared for a participant with a relatively high pneumococcal abundance (high piaB) and average 16S abundance (overall bacterial abundance), represented by the dashed curves and a participant with relatively low pneumococcal abundance (low *piaB*) and average 16S abundance, represented by continuous curves. The continuous curves decrease at a lower rate than the dotted counterparts, illustrating the adverse effect of high *piaB* values on the rate of infection acquisition. Curves from children and adults are depicted in black and grey colours, respectively.

**Figure S3:**
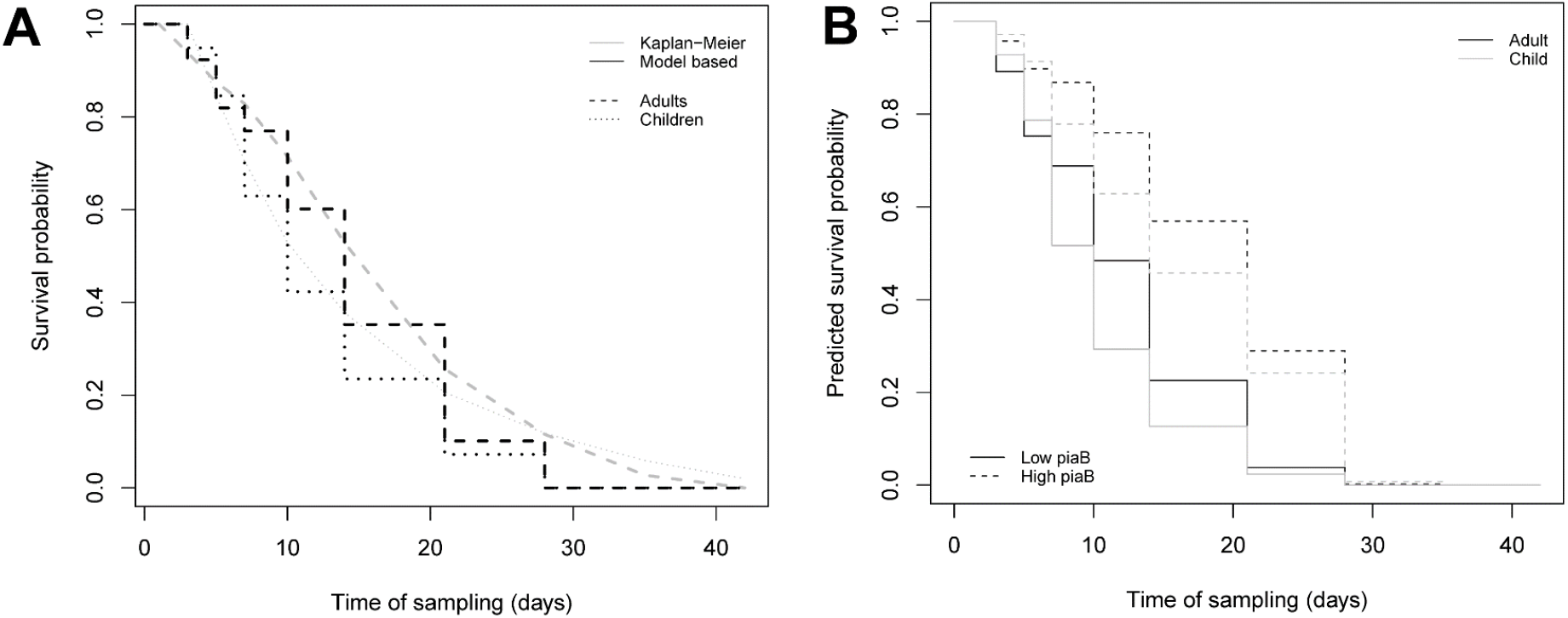
**(A)** The survival probability of time to infection acquisition estimated by the non-parametric Kaplan-Meier estimator (in grey) and model-based (in black) and the respective 95% confidence intervals. The two sets of curves are in agreement with each other, both for children and for adults. **(B)** Predicted survival probabilities based on the Cox proportional hazards model (model IV of Table S3), stratified by age group, were compared for a participant with a relatively high pneumococcal abundance (high *piaB*) and average 16S abundance (overall bacterial abundance), represented by the dashed curves and a participant with relatively low pneumococcal abundance (low piaB) and average 16S abundance, represented by continuous curves. The continuous curves decrease at a higher rate than the dotted counterparts, illustrating the impairing effect of high *piaB* values on SARS-CoV-2 infection clearance rates. Curves from children and adults are depicted with black and grey colours, respectively.

**Figure S4:**
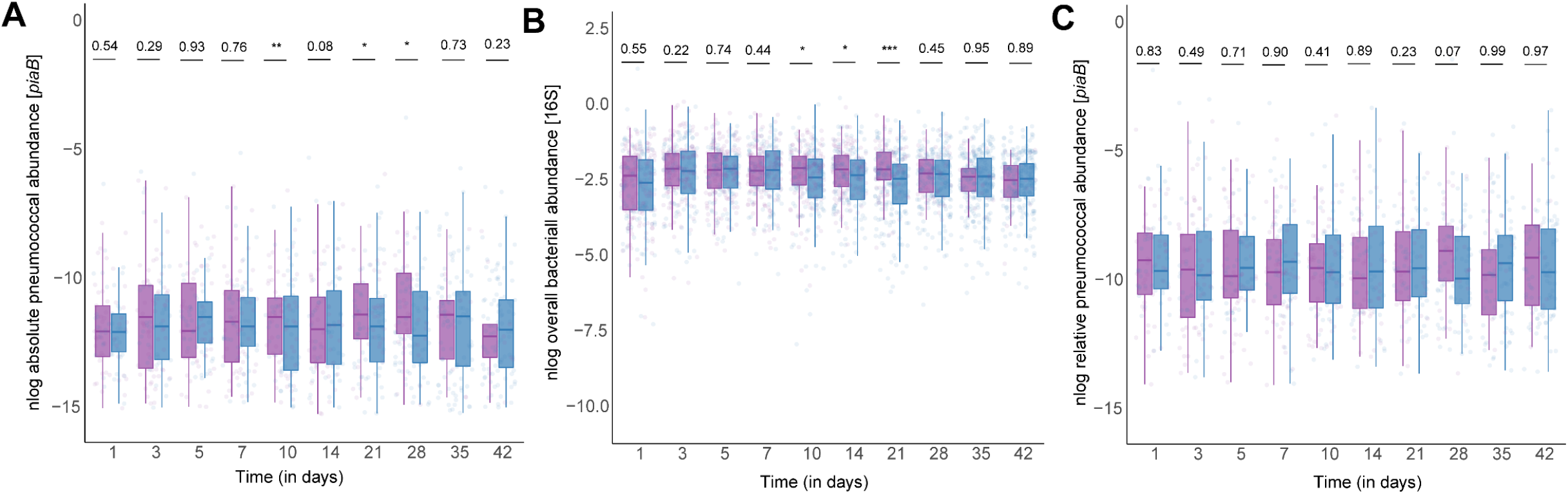
(A) Pneumococcal abundances (*piaB*) of individuals that are identified as carrier for ≥1 sampling events, stratified by COVID-19 status. (B) Overall bacterial abundances from any individual irrespective of pneumococcal carriage status, stratified by COVID-19 status. The colour purple indicates positivity for SARS-CoV-2 infection. None of the sampling events display a significant difference in either pneumococcal abundances or overall bacterial abundance between individuals with or without a SARS-CoV-2 infection. A permutation test equivalent of the Mann-Whitney U test was used to compare abundances of SARS-CoV-2 infected and uninfected individuals while blocking by age groups (< 18 years old or ≥ 18 years old).

**Figure S5:**
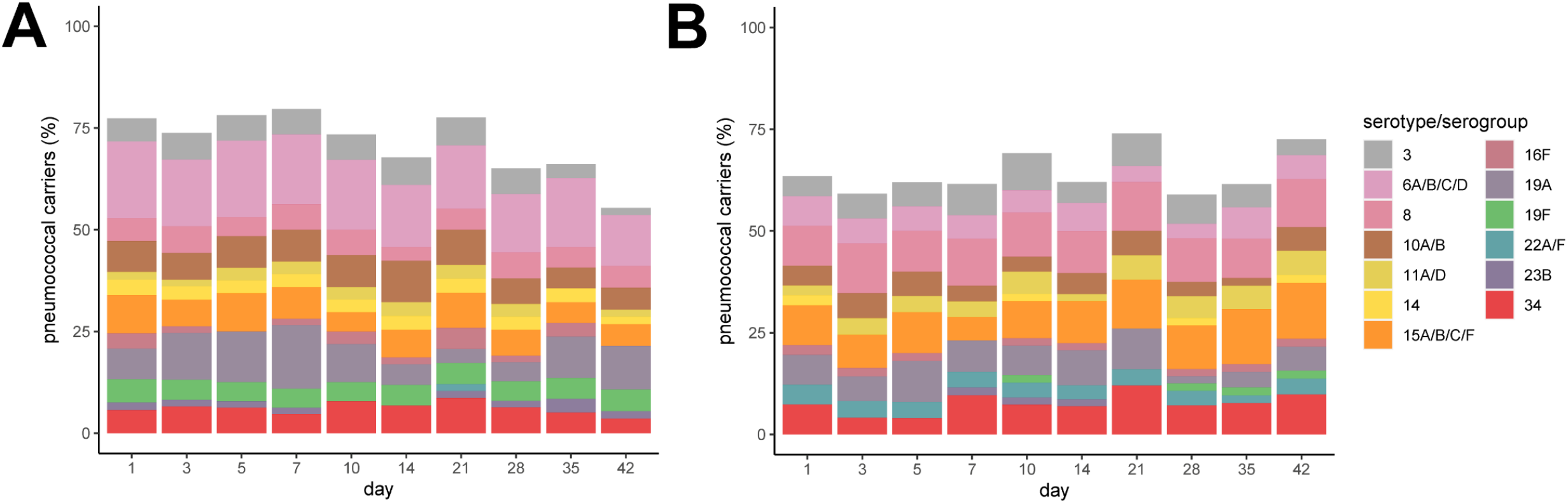
Stacked bardiagrams of serotype sample composition per time point for (A) children) and (B) adults. Analysis was limited to qPCR-targeted serotypes. The percentage of serotypes or serogroups among pneumococcus positive samples are shown. Percentages are calculated per sampling event for children (<18 year olds) and adults (≥ 18 year olds) separately. Results are limited to serotype-specific or serogroup-specific qPCR assays that produced atleast one or more positive samples.

**Figure S6:**
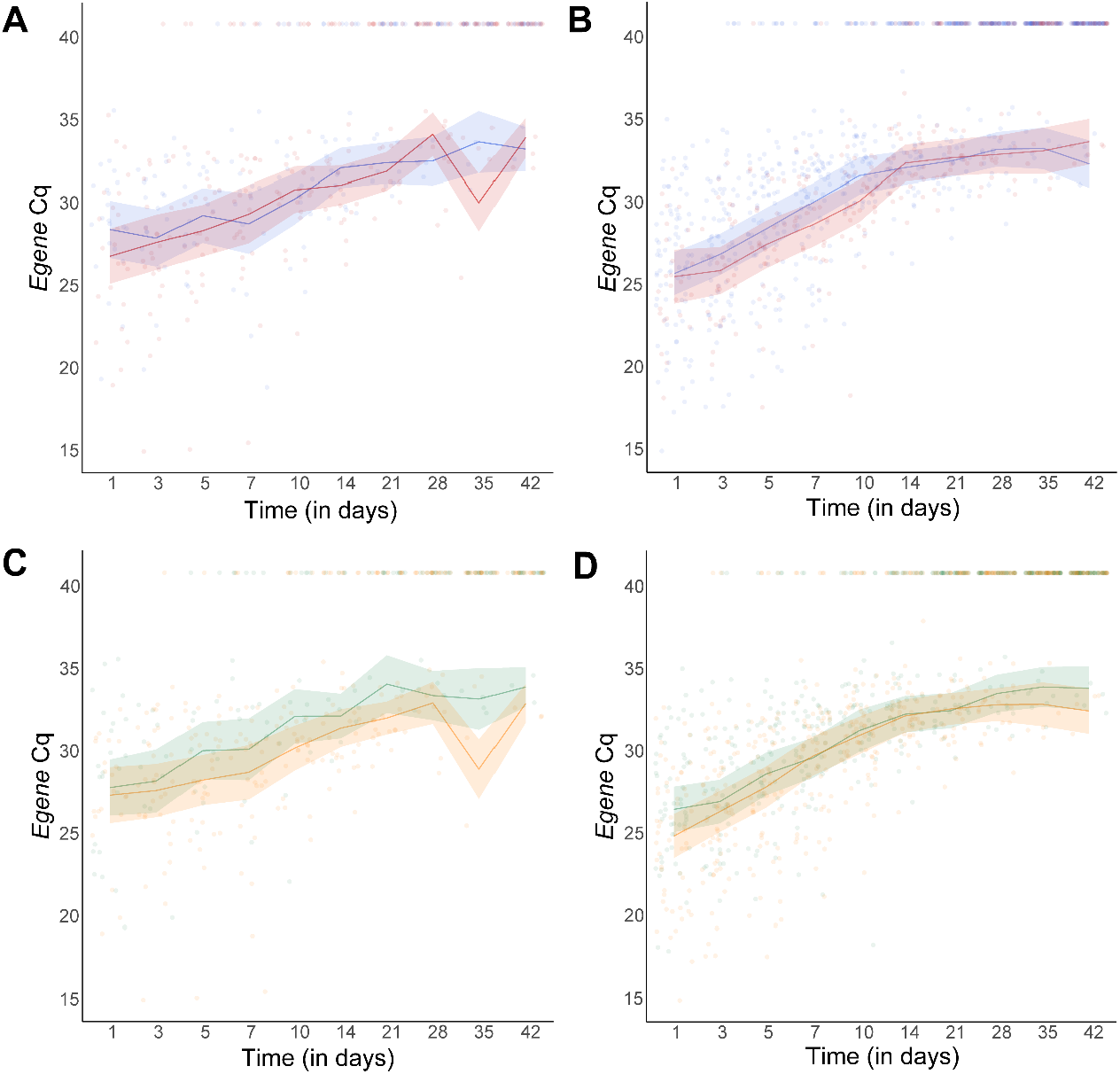
Longitudinal SARS-CoV-2 viral load trajectories of pneumococcal carriers (A & B), and individuals with 16S abundance (C & D) were analyzed with linear mixed effects models. Data from children (**A & C**) and adults (**B** & **D**) are modeled separately. Shaded areas indicate the 95% confidence interval of model estimates and the lines indicate the coefficients of model estimates. Censoring was applied to samples with a viral load of ≥ 40 Cq for the *Egene*. Pneumococcal carriers (A & B) are colored in red and noncarriers in blue. All individuals were positive for SARS-CoV-2 at T=1. Individuals with 16S abundances higher or equal to the median 16S abundance at T=1 are colored in green and individuals with 16S abundances below the median 16S abundance at T=1 are colored in yellow. None of the tested model exhibited significant differences in viral load trajectories for any of the analyzed groups.

## SUPPLEMENTAL TEXT

### Molecular Detection of Pneumococcal Serotypes

#### qPCR-based Serotyping of Saliva Samples

Per individual, DNA was extracted from pools of samples exclusively positive for *S. pneumoniae* and collected from a single person. For each individual, samples positive for *S. pneumoniae* were pooled in equal ratio and 200 *μ*l of the pool (for individuals positive for *S. pneumoniae* only once, 200 *μ*l of a single sample) was processed for nucleic acids extraction as described above. Serotype composition of pools was determined using a panel of primers and probes targeting serotypes 1, 3, 8, 14, 16F, 19A, 19F, 20, 21, 23A, 23B, 23F, 34 and 38, and serogroups 6A/B/C/D, 7A/F, 9A/L/N/V, 10A/B, 11A/D, 12A/B/C/F, 15A/B/C/F, 18A/B/C/F, 22A/F, 33A/F, 35B/C. When a pool of samples was positive for a serotype-specific or serogroup-specific qPCR assay all individual samples were also tested for that particular assay.

#### Evaluation of Diagnostic Specificity of Serotype/Serogroup-specific qPCR Assays in Saliva

Bland-Altman analysis was conducted to evaluate the specificity of serotype/serogroup-specific qPCR assays. Per sampling event, the upper limit of agreement between *piaB* and *lytA* was used as an a priori acceptible limit for agreement between serotype quantification and *piaB* or *lytA*. These limits ranged between 2.79 to 3.72 and were used to identify putative nonreliable results that exhibited strongly increased quantification of serotypes or serogroups when compared with quantification of *piaB* or *lytA*. In this analysis, serotypes 21 and 23A, and serogroups 9A/L/N/V, 12A/B/F, 33A/F and 35B/C were overrepresented and therefore all results from these assays were excluded from further analysis. Prior to exclusion of these qPCR assays, the mean ICC between dominantly ranked serotypes and *piaB* was 0.61, whereas after exclusion the mean ICC was 0.70.

## Notes

### Competing Interest Statement

We report that K. Trzcinski has received consultation and speaking fees and funds for unrestricted research grants from Pfizer, funds for unrestricted research grants from GlaxoSmithKline, and consultation fees from Merck Sharp & Dohme, all paid directly to his home institution. None of these funds contributed to the present study. All other authors report no potential conflicts of interest.

### Funding Statement

This study did not receive any funding.

### Author Declarations

Written informed consent was obtained from all study participants or from their legal guardians. The study was reviewed and approved by the Medical Ethical Committee of the Vrije Universiteit university Medical Centre (Vumc), The Netherlands (reference nr. 020.436).

